# Investigating brain dynamics and their association with cognitive control in opioid use disorder using naturalistic and drug cue paradigms

**DOI:** 10.1101/2024.02.25.24303340

**Authors:** Jean Ye, Saloni Mehta, Hannah Peterson, Ahmad Ibrahim, Gul Saeed, Sarah Linsky, Iouri Kreinin, Sui Tsang, Uzoji Nwanaji-Enwerem, Anthony Raso, Jagriti Arora, Fuyuze Tokoglu, Sarah W. Yip, C. Alice Hahn, Cheryl Lacadie, Abigail S. Greene, R. Todd Constable, Declan T. Barry, Nancy S. Redeker, Henry Yaggi, Dustin Scheinost

**Affiliations:** Interdepartmental Neuroscience Program, Yale University; Department of Radiology & Biomedical Imaging, Yale School of Medicine; Department of Health Policy, Vanderbilt University; Department of Internal Medicine, Yale School of Medicine; Department of Internal Medicine, Roger Williams Medical Center; Yale School of Nursing; Pulmonary, Critical Care and Sleep Medicine, Yale School of Medicine; Program of Aging, Yale University; Frank H. Netter M.D. School of Medicine, Quinnipiac University; Department of Psychiatry, Yale School of Medicine; Child Study Center, Yale School of Medicine; Yale Center for Clinical Investigation, Yale School of Medicine; Department of Psychiatry, Brigham and Women’s Hospital; Department of Biomedical Engineering, Yale School of Engineering and Applied Science; Department of Neurosurgery, Yale School of Medicine; Department of Research, APT foundation; School of Nursing, University of Connecticut; Clinical Epidemiology Research Center, VA CT Healthcare System; Department of Statistics & Data Science, Yale School of Medicine

## Abstract

**Objectives:** Opioid use disorder (OUD) impacts millions of people worldwide. The prevalence and debilitating effects of OUD present a pressing need to understand its neural mechanisms to provide more targeted interventions. Prior studies have linked altered functioning in large-scale brain networks with clinical symptoms and outcomes in OUD. However, these investigations often do not consider how brain responses change over time. Time-varying brain network engagement can convey clinically relevant information not captured by static brain measures.

**Methods:** We investigated brain dynamic alterations in individuals with OUD by applying a new multivariate computational framework to movie-watching (i.e., naturalistic; N=76) and task-based (N=70) fMRI. We further probed the associations between cognitive control and brain dynamics during a separate drug cue paradigm in individuals with OUD.

**Results:** Compared to healthy controls (N=97), individuals with OUD showed decreased variability in the engagement of recurring brain states during movie-watching. We also found that worse cognitive control was linked to decreased variability during the rest period when no opioid-related stimuli were present.

**Conclusions:** These findings suggest that individuals with OUD may experience greater difficulty in effectively engaging brain networks in response to evolving internal or external demands. Such inflexibility may contribute to aberrant response inhibition and biased attention toward opioid-related stimuli, two hallmark characteristics of OUD. By incorporating temporal information, the current study introduces novel information about how brain dynamics are altered in individuals with OUD and their behavioral implications.

## Introduction

Opioid use disorder (OUD) is a chronic condition impacting millions of people globally (1). Individuals with OUD face increased risks of injuries, incarceration, and premature mortality (2,3). Given OUD’s profound burden on the individual and the community, there is an urgent need to elucidate its etiology to develop more targeted preventions and interventions (4).

Recent work using functional magnetic resonance imaging (fMRI) has greatly expanded our understanding of OUD (5–7). Task-based fMRI collected during drug cue and response inhibition paradigms has been widely utilized to investigate OUD’s neural mechanisms. These studies implicated the involvement of several canonical functional brain networks, including the default mode (DMN), reward, and cognitive control networks (5,8–10). Specifically, individuals with OUD expressed hyperactivation in the striatum, amygdala, and dorsolateral prefrontal cortex (PFC) in response to opioid-related stimuli (7,11). Anterior cingulate and lateral and medial PFC hypoactivation were additionally observed in individuals with OUD when the need to suppress automatic response arose (7,12,13).

However, extant fMRI studies adopt a static approach, where brain activity is averaged over a minutes-long scan. Therefore, these methods are unable to track brain activation fluctuations across time. How individuals dynamically engage brain networks contains crucial information about psychopathology. Notably, dynamic brain markers reveal insights about clinical symptoms not captured by static brain measures computed from the average activity patterns over a single scan (14,15). Thus, not considering the dynamic changes in brain activity over time may contribute to an incomplete understanding of OUD.

To address this gap, we leveraged a new multivariate computational framework to investigate brain dynamics in individuals with OUD stabilized on medication for OUD (MOUD). Our framework has the advantage of simultaneously tracking multiple brain states (i.e., recurring brain activation patterns) at the resolution of individual time points. As brain states can overlap temporally (16,17), examining multiple brain states concurrently addresses the caveat of potentially neglecting less dominant brain states with important behavioral relevance. A helpful analogy to consider here is color (**Figure 1**). If blue (representing the most dominant brain state) contributes the most to purple (representing a time point of interest) and only blue is examined, information on how other colors like red (representing the less dominant brain states) also contribute to forming purple is lost. This framework allows us to assess variability in brain state engagement over time by extracting moment-to-moment engagement information for multiple brain states simultaneously.

**Figure 1.**
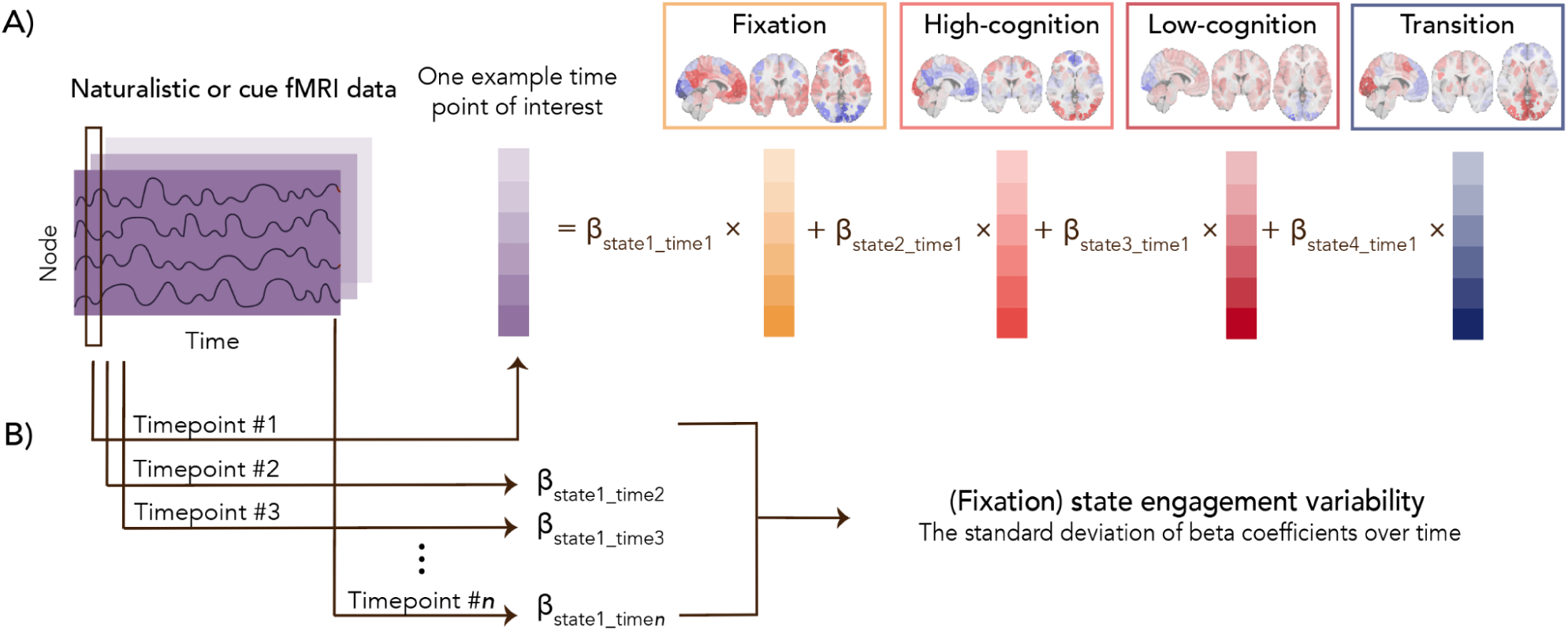
Study pipeline. Four recurring brain states were identified using task-based fMRI from the Human Connectome Project dataset. Based on the prominent task conditions associated with each brain state, we characterized these four brain states as fixation, high-cognition, low-cognition, and transition (**Supplementary Materials**). For example, the fixation brain state mainly includes time points from the fixation condition across task paradigms. In contrast, the low-cognition brain state consists of time points from the motor task, the 0-back working memory task condition, and the neutral emotion task condition (**Supplementary Table 1**). Moment-to-moment state engagement was assessed in the naturalistic and drug cue paradigms using non-negative least squares regression. For a given time point, the framework can assess the engagement (indicated by the beta coefficients) of multiple brain states concurrently. **A** demonstrates this process for one example time point of interest. After engagement values are extracted from all time points, variability is computed as the standard deviation of moment-to-moment engagement across time. As an example, **B** shows how variability is computed for the fixation state.

Using fMRI data collected during a movie-watching paradigm, we first compared brain state engagement variability between individuals with OUD and healthy controls (HCs). Naturalistic stimuli closely mimic the time-varying interactions in real life (18), potentially presenting a more ecologically valid understanding of brain dynamics in OUD. Increased brain network recruitment variability supports greater flexibility in addressing current demands (19,20). As decreased cognitive flexibility has been observed in individuals with OUD (21,22), we hypothesized that they would show decreased state engagement variability compared to HCs during movie-watching. Next, taking advantage of task-based fMRI’s increased power in detecting brain-behavior associations (23), we correlated brain dynamics during a drug cue paradigm with Stroop-assessed cognitive control. We hypothesized that lower variability in the drug cue paradigm, particularly during rest block, would be associated with worse cognitive control.

Additionally, despite notable sex differences in the mortality rate and risk for OUD (24,25), an understanding of sex differences in OUD at the neurobiological level is lacking (7). To address this gap, we explored sex effects on brain dynamics and their associations with behaviors.

## Methods

### Participants

Data from two datasets were analyzed. The Yale University Institutional Review Board and the Yale MRRC Protocol Review committee approved both studies. Informed consent was obtained from all participants. The OUD sample was recruited as a part of an NIH-funded study from Yale School of Medicine and the APT foundation (Collaboration Linking Opioid Use Disorder and Sleep). It included individuals who met DSM-5 criteria for OUD and who were stabilized on MOUD (<24 weeks), an evidence-based treatment for OUD. Naturalistic and drug cue fMRI data were examined here. During the 6-minute naturalistic paradigm, participants watched three movie clips presented in the same order without a break (*Inside Out*, *The Princess Bride*, and *Up)*. In the drug cue paradigm, participants were presented with either a rest (a white crosshair presented in the center of a black screen) or a cue block (a picture showing opioid-related stimuli; e.g., a needle or a bottle of pills). There were nine alternating rest and cue blocks, each lasting 16 seconds.

HCs from a separate transdiagnostic study were also studied. Participants were only included if they had no neurological or mental health diagnoses based on the Mini-International Neuropsychiatric Interview for DSM-5. Naturalistic fMRI data were collected using the same paradigm described above. No drug cue data were collected from HCs.

### FMRI data preprocessing

Detailed acquisition and preprocessing information can be found in **Supplementary Materials**. In brief, a standard preprocessing pipeline described in previous studies was applied to the structural and functional data. Statistical Parametric Mapping (SPM12) performed slice time and motion correction for both datasets on the functional data. Additional data cleaning was carried out in BioImage Suite.

Covariates of no interest were regressed out, including linear and quadratic drift, white matter, cerebrospinal fluid, gray matter, and a 24-parameter motion model. The functional data were temporally smoothed (cutoff frequency around 0.12Hz). A combination of nonlinear and linear transformations aligned the Shen-268 atlas to the functional data before timeseries data was parcellated.

Additional quality control included removing participants with mean framewise displacement over 0.2mm, missing time points, or brain coverage (see **Supplementary Materials** for more details). After these criteria, 97 HCs (**Table 1**) and 76 individuals with OUD (**Table 1**) were included in the naturalistic fMRI analysis; 70 individuals with OUD with drug cue fMRI data (**Table 1**) were analyzed.

**Table 1.** Demographic information for both datasets. OUD duration was computed as the difference between their current and age at first use. OUD severity was measured using SCID-V based on DSM-5 (mild: 2-3 symptoms, moderate: 4-5 symptoms, severe: 6 or more symptoms). HC, healthy control. OUD, individuals with opioid use disorder.

|  | Characteristic |  |  |
| --- | --- | --- | --- |
| Naturalistic paradigm (HC) | N |  | 97 |
|  | Self-reported sex assigned at birth | Female | 54 |
|  |  | Male | 43 |
|  | Age (years) | Mean | 31.660 |
|  |  | Standard deviation | 12.178 |
|  | Self-reported race | Asian | 16 |
|  |  | Black | 23 |
|  |  | More than one race | 5 |
|  |  | Unknown | 1 |
|  |  | White | 52 |
| Naturalistic paradigm (OUD) | N |  | 76 |
|  | Self-reported sex assigned at birth | Female | 31 |
|  |  | Male | 45 |
|  | Self-reported race | Asian | 0 |
|  |  | Black | 2 |
|  |  | More than one race | 6 |
|  |  | Unknown | 3 |
|  |  | White | 65 |
|  | OUD duration | Mean | 14.816 |
|  |  | Standard deviation | 9.927 |
|  | OUD severity | Moderate | 4 |
|  |  | Severe | 72 |
|  | Methadone dose at baseline (milligram) | Mean | 81.947 |
|  |  | Standard deviation | 22.644 |
| Drug cue paradigm | N |  | 70 |
|  | Self-reported sex assigned at birth | Female | 31 |
|  |  | Male | 39 |
|  | Age (years) | Mean | 39.2 |
|  |  | Standard deviation | 10.691 |
|  | Self-reported race | Asian | 0 |
|  |  | Black | 2 |
|  |  | More than one race | 6 |
|  |  | Unknown | 3 |
|  |  | White | 59 |
|  | OUD duration | Mean | 14.686 |
|  |  | Standard deviation | 9.909 |
|  | OUD severity | Moderate | 3 |
|  |  | Severe | 67 |
|  | Methadone dose at baseline (milligram) | Mean | 81.5 |
|  |  | Standard deviation | 22.847 |

### Brain dynamic measures

We leveraged a new multivariate computational framework to assess brain dynamics at the whole-brain level (26; **Supplementary Materials; Figure 1**). Briefly, four recurring brain states were identified by applying nonlinear manifold learning to task-based fMRI data from the Human Connectome Project (HCP) dataset (27). These brain states were later characterized as fixation, high-cognition, low-cognition, and transition based on the task conditions and their associated cognitive load. For instance, the time points in the high-cognition brain state are from complex cognitive paradigms requiring higher cognitive load (e.g., working memory, gambling, and emotion). The transition brain state consists of time points when individuals switched from one task condition to another. The fixation brain state includes time points where individuals were presented with a crosshair while resting in the scanner (**Supplementary Table 1**). The rich HCP dataset allowed us to identify brain states underlying a range of cognitive processes while circumventing circular analysis. While HCP consists of healthy individuals, previous work has indicated that similar brain states can be identified in individuals with psychopathology, suggesting that the way brain states are recruited, instead of the brain states themselves, is more likely to drive group differences (17,28,29).

Non-negative least squares regression next extended these four brain states to our two datasets, evaluating each state’s engagement at each time point during both the naturalistic and drug cue paradigms. Our approach permits temporal overlap in brain states and allows for the joint contributions of all brain states to a given time point. This flexibility is crucial, as the activation pattern observed at a single time point might not always match exactly onto a specific brain state. A summary measure was then extracted for each individual and each brain state by computing the standard deviation of moment-to-moment engagement across time (i.e., state engagement variability). Variability was calculated across the entire run for the naturalistic paradigm. As prior work has revealed significant differences in variability during rest and task (26), we opted to compute variability within the rest and cue condition separately for the drug cue paradigm. As a validation analysis, we compared state engagement variability between conditions in the drug cue paradigm to test whether our brain dynamic measures were sensitive to changes induced by task demands. The more constrained nature of the cue condition likely reduces the likelihood of mind wandering. If our measures are sensitive to brain dynamic changes in response to variations in demand, we may observe lower variability during cue in contrast to the rest condition.

### Stroop-assessed cognitive control

We extracted cognitive control measures from a Stroop task to evaluate the behavioral implications of alterations in state engagement variability. Both accuracy (ACC; i.e., control trial ACC - incongruent trial ACC) and response time (RT; i.e., incongruent trial RT - control trial RT) interference scores were computed. Individuals with missing or outlier behavioral scores were excluded. We included 62 and 61 participants with OUD in the final ACC and RT interference score analysis (see exclusion criteria in **Supplementary Materials**), respectively.

### Statistical Analysis

Multivariate group differences in state engagement variability during movie-watching were examined with Hotelling’s T-square test. Age and sex were included as covariates. Sex-by-group and age-by-group interactions were explored. We additionally correlated Stroop interference scores with state engagement variability during the drug cue paradigm using Spearman correlation. There may be lasting effects of opioid-related stimuli in the rest block, thereby making this period more sensitive to flexibility in the brain. To probe this possibility, the first rest block was excluded when computing variability for consistency since no prior stimuli were presented. We also examined the association between cognitive control and state engagement variability during the cue condition for completeness. Results for these analyses were FDR corrected for multiple comparisons.

## Results

### Demographic data

Demographic information for healthy controls and individuals with OUD is included in **Table 1**. All individuals with OUD were stabilized on methadone and met DSM-5 criteria for either moderate or severe OUD (see **Supplementary Table 3** for additional clinical characteristics). Among individuals included in the naturalistic paradigm analyses, HCs were significantly younger than individuals with OUD (two-sample t-test; t(169.49)=-4.472; p<0.001). The two groups did not differ in sex (Chi-squared; p=0.073).

### Individuals with OUD demonstrated lower state engagement variability

We observed a significant group effect on state engagement variability during movie-watching (F(4,164)=9.967; p<0.001; **Figure 2**) when including age and sex as covariates. There was no sex main effect (F(4,164)=1.323; p=0.264) or sex-by-group interaction (F(4,164)=0.327, p=0.860). Consistent with previous work (26), age showed a significant effect on variability (F(4,164)=6.329, p<0.001), but there was no age-by-group interaction (F(4,164)=2.295, p=0.061).

**Figure 2.**
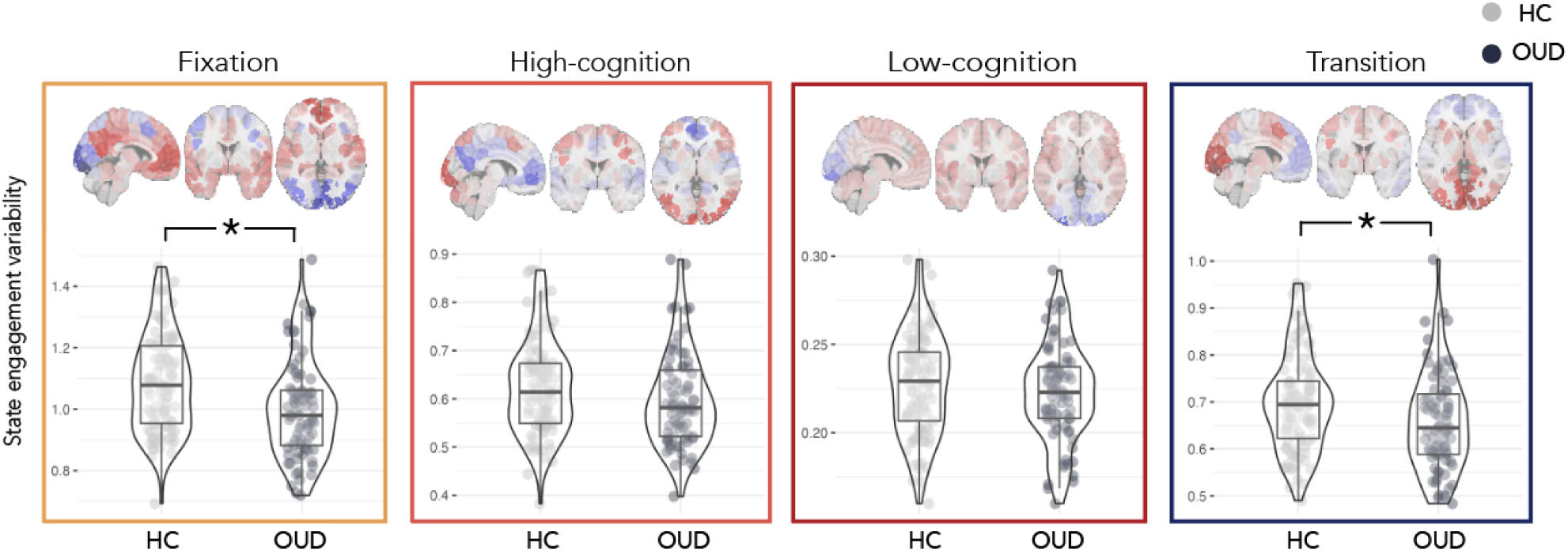
Group differences in state engagement variability. During naturalistic fMRI, individuals with OUD (N=76) showed lower state engagement variability than HCs (N=97). This was prominent in the fixation and transition brain states. OUD, opioid use disorder; HC, healthy control. Scatter plots created using the ggstatsplot R package (30).

Post-hoc ANOVAs were performed to explore group differences in each brain state separately (**Supplementary Table 4**). Fixation (F(1,167)=12.343; p<0.001) and transition state engagement variability (F(1,167)=6.667; p=0.011) were significantly lower in individuals with OUD compared to HCs (fixation: HC: 1.085±0.159, OUD: 1.000±0.160; transition: HC: 0.698±0.103, OUD: 0.658±0.104).

High-cognition (F(1,167)=2.455; p=0.119) and low-cognition state engagement variability (F(1,167)=1.270; p=0.261) did not differ significantly between groups.

### Lower state engagement variability during rest condition was associated with worse cognitive control

As expected, our validation analysis revealed that state engagement variability decreased during cue compared to rest across all four states (fixation: t(69)=2.014, p=0.048; high-cognition: t(69)=2.104, p=0.039; low-cognition: t(69)=4.623, p<0.001; transition: t(69)=2.136, p=0.036). These patterns remained consistent even when we introduced lags to task time indices (**Supplementary Table 5**), indicating that our measures are sensitive to brain dynamic changes following demand changes in the environment.

Notably, while state engagement variability during the cue condition was not linked to cognitive control (i.e., ACC or RT interference scores; **Table 2**), cognitive control was significantly correlated with variability assessed during the rest condition. Specifically, decreased transition and low-cognition state engagement variability was linked to worse ACC (ρ(60)=-0.405; p=0.001; q=0.016) and RT interference (ρ(59)=-0.383; p=0.002; q=0.016), respectively (**Figure 3**; **Table 3**). These associations remained largely consistent when lags were added to task time indices (**Supplementary Tables 6 & 7**).

**Figure 3.**
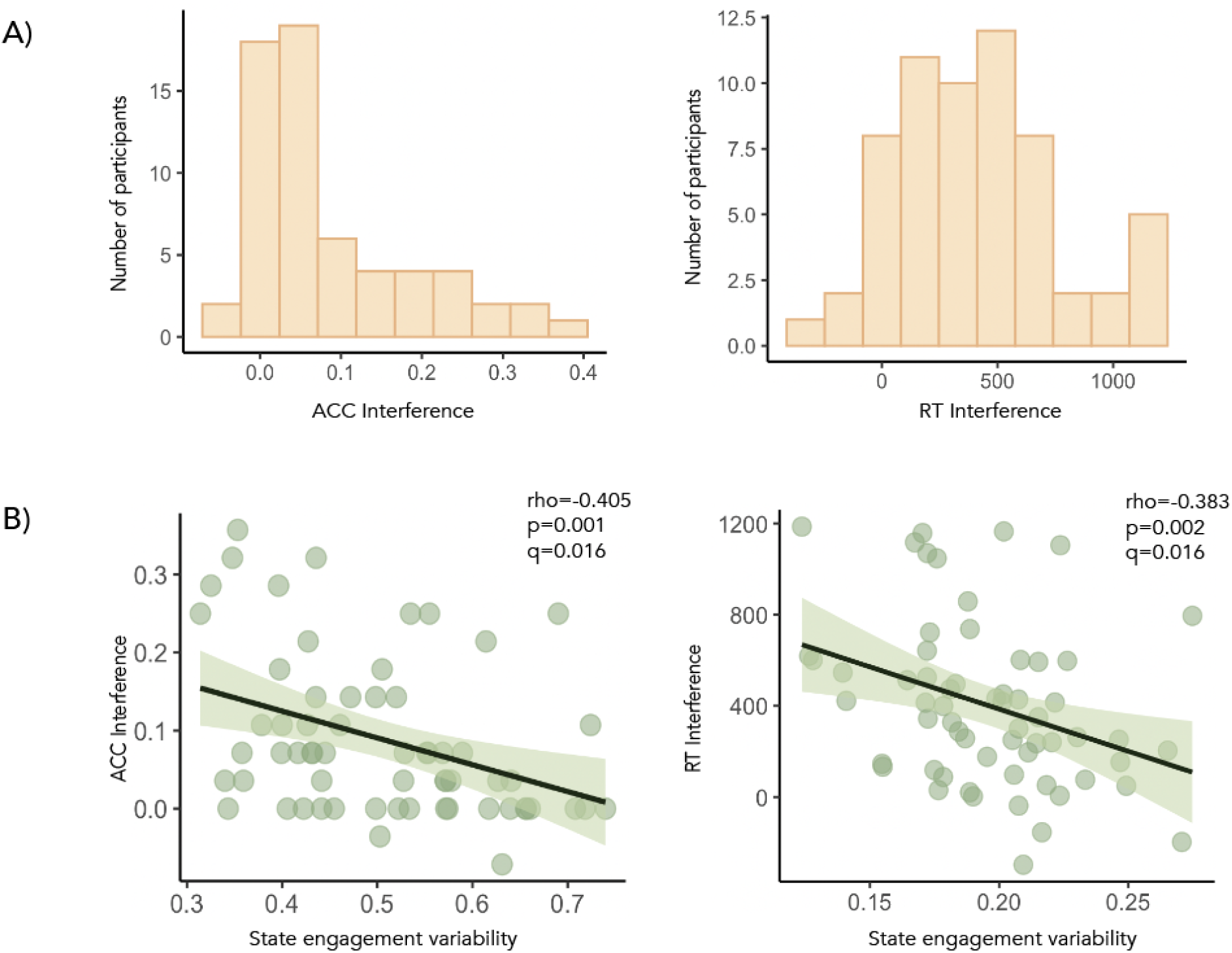
State engagement variability and cognitive control. ACC and RT interference score distributions were shown in **A**. **B** During the rest condition of the drug cue paradigm, decreased transition state engagement variability and low-cognition state engagement variability were associated with worse cognitive control performance (i.e., higher ACC and RT interference scores), respectively.

**Table 2.**
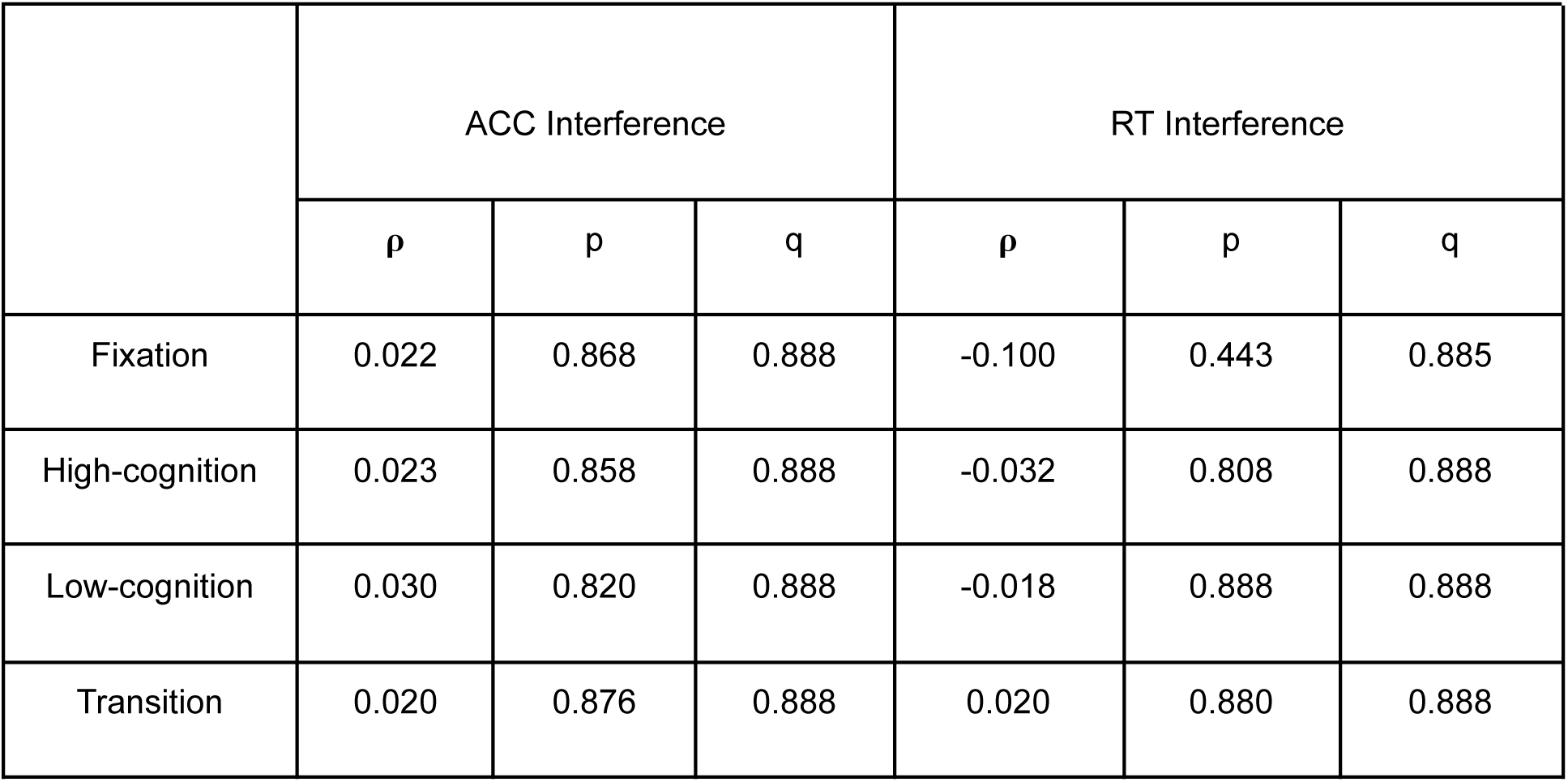
State engagement variability during cue and cognitive control.

**Table 3.**
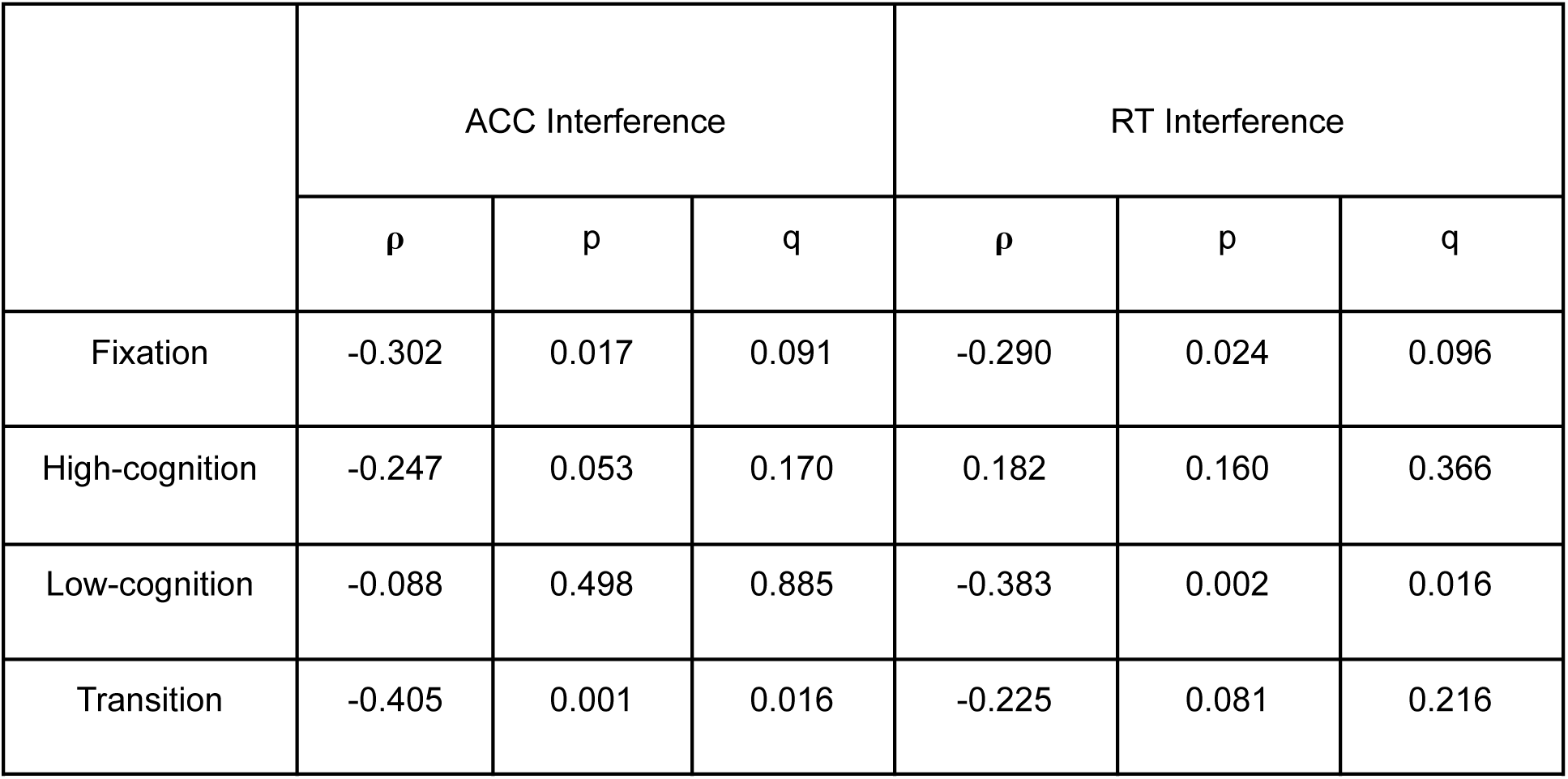
State engagement variability during rest and cognitive control.

Exploratory analysis was conducted to investigate whether these significant associations varied by sex. Interestingly, ACC interference was only significantly correlated with transition state engagement variability in male (ρ(30)=-0.516; p=0.003) but not female participants (ρ(28)=-0.228; p=0.226). In contrast, RT interference was significantly linked to low-cognition state engagement variability in female (ρ(29)=-0.580; p<0.001) but not male participants (ρ(28)=-0.174; p=0.357). Group comparison of these correlations were not significant (two-tailed; ACC: z=1.22, p=0.223; RT: z=-1.74, p=0.082).

### Control analyses

Our framework assessed variability in the moment-to-moment engagement of recurring whole-brain brain states. As a control analysis, we extracted variability in brain node activation to investigate whether similar results can be obtained (31; **Supplementary Materials**). Briefly, node activation variability is computed as the average of variability in activation timeseries across all brain nodes. Node activation variability did not differ significantly between groups (ANOVA; F(1,180)=1.602, p=0.207) or correlate with ACC interference scores (rest node activation variability: ρ=-0.084, p=0.517; cue node activation variability: ρ=0.044, p=0.735). RT interference scores correlated significantly with node activation variability during rest (ρ=-0.349, p=0.006) and cue (ρ=-0.279, p=0.030). These preliminary results indicate that compared to variability in node activation across time, our state engagement variability measures may be more sensitive to detecting group differences in brain dynamics. While significant associations between variability and RT interference aligned with previous literature (32,33), the effect sizes found here were smaller than those observed with our brain dynamic measures.

We additionally explored whether brain responses during the drug cue paradigm were related to cognitive control. A general linear model analysis was performed (**Supplementary Materials; Supplementary Figure 1**). Beta coefficients were extracted from each brain node and correlated with ACC and RT interference scores. After correction for multiple comparisons, none of the brain nodes demonstrated responses significantly correlated with cognitive control (**Supplementary Table 8**).

## Discussion

We leveraged a new multivariate computational framework to investigate alterations in brain dynamics in individuals receiving MOUD for OUD. During movie-watching, individuals with OUD demonstrated reduced state engagement variability compared to HCs. Decreased variability during the rest condition of a drug cue paradigm was further associated with worse cognitive control in individuals with OUD. The novelty of the current study is twofold. First, it served as an initial investigation of brain dynamic alterations in OUD. By employing naturalistic fMRI in our exploration, we built a more ecologically valid understanding of brain dynamics and contributed new insights about OUD. Second, lower variability in brain state engagement was linked to worse cognitive control. By analyzing brain activity across time, we found that a task classically used to study one hallmark characteristic of OUD (i.e., aberrant response to opioid-related stimuli) could provide information on another symptom (i.e., impaired cognitive control). Notably, a traditional brain activation analysis did not reveal an association between cognitive control and brain responses during the drug cue paradigm.

Broadly, state engagement variability is a form of neural variability. Once considered noise, neural variability is now recognized to support behavioral flexibility. Increased variability allows the brain to flexibly recruit various brain networks to call on different processes in response to current needs (19,20,34). Thus, decreased state engagement variability may reflect an impairment in effectively engaging different brain states to address evolving demands. By incorporating time information, our approach provides information about how network recruitment over time may be different in OUD.

The interpretation that altered variability reflects disrupted brain network communications in OUD aligns with the reduced structural and functional brain network connections found in mice and humans dependent on opioids (35,36). We observed significantly reduced variability in the fixation and transition brain states in individuals with OUD. The fixation brain state predominantly recruits the DMN (**Supplementary Table 2**), which is relevant for relapse risk and withdrawal symptoms in individuals using heroin (37,38). The transition brain state captures activation patterns that often appear when individuals respond to a change in task demand (**Supplementary Table 1**). Taken together, the inability to effectively recruit these two brain states may lead to greater difficulty switching away from self-relevant thoughts supported by the DMN. This possibility can have important clinical implications if these self-related thoughts revolve around the urge or motivation for opioid use (e.g., the physical or mental pain one is experiencing). These perseverative thoughts may further contribute to craving or substance-seeking behaviors.

Our findings in the naturalistic paradigm indicated that lower state engagement variability might be a characteristic of OUD. Further investigations with variability in a drug cue paradigm demonstrated that this decreased variability could have important behavioral relevance. Consistent with prior literature suggesting that neural variability supports behavioral performance (32,33,39), we found that decreased state engagement variability was associated with worse cognitive control in individuals with OUD. An elevated ACC interference score was linked to lower transition engagement variability, suggesting that the flexible recruitment of a brain state relevant to task demand change supports cognitive control. Further, a higher RT interference score was linked to decreased low-cognition state engagement variability. The low-cognition brain state recruits the motor and medial frontal networks (**Supplementary Table 2**). Notably, the activity of these two networks during a Stroop task predicts opioid abstinence (6). These findings indicate that more effective engagement of these brain networks may contribute to better cognitive control over the automatic or habitual motor-based responses associated with substance-seeking behaviors (40,41). Future research can probe whether the flexible engagement of the low-cognition brain state relates to abstinence and whether individual variations in cognitive control may mediate this relationship.

Interestingly, significant associations between cognitive control and state engagement variability were found in the rest but not cue condition. This finding dovetails with previous work suggesting that—in addition to the initial response to salient stimuli (such as opioid-related images)—the lingering or recovery from such stimuli also conveys important information (42–44). Drug cue paradigms are a popular approach to investigate the neural mechanisms underpinning OUD. While brain responses to opioid-related stimuli relate to treatment adherence and withdrawal symptoms (45,46), less work has focused on the rest period with no cue presentation. Our results indicate that this period may serve as a novel window into studying stickiness (i.e., residual engagement or decreased flexibility) in the brain. The lower variability observed during naturalistic stimuli in individuals with OUD might reflect more restricted brain network recruitment (19) and a general tendency to linger in a particular thought pattern. This characteristic may become even more prominent in the rest period after opioid-related stimuli are presented. Interestingly, variability during the rest period before presentation of opioid-related stimuli was not related to cognitive control (**Supplementary Table 9**). Altogether, these results suggest that following exposure to opioid-related stimuli, brain responses to these stimuli may linger due to decreased variability, potentially contributing to perseverative thoughts about opioids and inducing stronger cravings.

However, lingering opioid-related processing is only one of many interpretations of what lower variability during rest reflects. Rest also appears before a cue block. Instead of altered inhibition leading to more persistent processing, biased attention towards opioid-related stimuli may also constrain thoughts and reduce variability during rest. Both response inhibition and biased attention for substance-related stimuli are hallmark characteristics of OUD (7,8,10,47,48) and substance use disorders (49,50). Since we did not instruct participants to disengage from the opioid-related pictures they saw, it is unclear whether there was an actual attempt to inhibit such thoughts during rest. We additionally did not assess how much perseveration participants experienced. Thus, understanding what contributes to lower variability during rest and whether it is linked to more intense craving or drive for substance-seeking behaviors remains a future research direction.

Sex is an important factor in OUD. We did not find significant sex-by-group interaction on state engagement variability during movie watching. However, the strength of the associations between cognitive control and variability varied by sex. While our sample size allows us to study sex differences, power was likely low. Further work on sex differences in brain dynamics in OUD is urgently needed.

Opioid use and overdose deaths increase at a more rapid rate in women than men (25). A faster progression to substance misuse has also been consistently observed in women (51). The possibility that similar brain dynamic alterations may contribute to varying behavioral profiles in different groups is critical to consider. Future studies investigating this topic can contribute to more targeted interventions.

The current study also has several limitations. Individuals with OUD present a heterogeneous sample, with remarkable interindividual variations in risk, susceptibility, disorder profile, and progression. More understanding of how comorbidities or polysubstance use may interact with opioid use to influence brain dynamics is needed. Additionally, all participants analyzed in this study were on methadone treatment. As there are multiple treatment options available for OUD (52), future investigations should explore whether brain dynamics may be altered in similar ways across individuals with OUD on different medications (e.g., buprenorphine, extended-release naltrexone) or at different stages of intervention.

Abstinence time, treatment time, and time since last dose have all been shown to affect brain responses to cues and cognitive control performance in individuals with OUD (46,53,54). Future work should examine whether different time scales interact with each other. For instance, would individuals abstinent for longer (i.e., relatively longer time scale) show different brain dynamics alterations (i.e., relatively shorter time scale)? Future studies can additionally select naturalistic stimuli specific to substance-use (e.g., scenes of opioid use) to probe brain dynamic alterations more precisely in OUD. Furthermore, as the OUD sample in this study consists predominantly of white individuals, additional research in a more diverse sample is needed to replicate the findings here.

In summary, applying a multivariate framework to naturalistic and task-based fMRI data unveiled significant alterations in brain dynamics in individuals with OUD compared to HCs. This aberrant ability to engage brain networks flexibly and effectively over time can have implications for cognitive control, which may be particularly important to consider during exposure to opioid-related stimuli. The current study presented a novel understanding of altered brain dynamics in OUD and their behavioral consequences.

## Data Availability

Data supporting findings of this study are available from the authors upon reasonable request.

## Acknowledgements

We would like to thank the participants who took part in the study.

## Supplementary Materials

### Functional magnetic resonance imaging (fMRI) acquisition and preprocessing

FMRI acquisition parameters were the same in both datasets and have been detailed in previous work (1). FMRI data were collected with harmonized Siemens 3T scanners using a 64-channel head coil at Yale’s Magnetic Resonance Research Center. An anatomical scan was collected using a magnetization-prepared rapid gradient echo sequence (repetition time=2400ms, echo time=1.22ms, voxel size=1×1×1mm). FMRI was acquired using a multiband gradient echo-planar imaging sequence (repetition time=1000ms, echo time=30ms, voxel size=2×2×2mm, multiband factor=5). Participants watched three movie clips for the naturalistic paradigm without a break (*Inside Out*, *The Princess Bride*, and *Up*). In the drug cue paradigm, participants were presented with alternating rest and cue blocks. Each condition had 9 blocks, each lasting 16 seconds. Participants were shown either a fixation crosshair (rest condition) or a picture depicting opioid-related stimuli (cue condition).

Neuroimaging data were collected from 103 participants with OUD. For the naturalistic paradigm, we first excluded participants who did not complete the task or had mean framewise displacement (MFD) over 0.2mm (N=24). One participant was excluded due to having epilepsy. After removing two additional participants with missing brain coverage, a final sample of 76 individuals with OUD was analyzed for group comparison. For the drug cue task, we removed 29 participants who did not complete the drug cue task or showed excessive motion (i.e., MFD > 0.2mm). Two participants were excluded due to missing time points in their scan. Two participants were removed due to missing brain coverage. After these exclusion criteria, we extracted brain dynamic measures from 70 participants with OUD.

The transdiagnostic study collected neuroimaging data from 307 participants. Out of these individuals, 294 adult participants completed the naturalistic paradigm. We excluded one participant with missing brain coverage. Three participants were further excluded due to having a wrong number of volumes. Of the 290 participants, 114 were considered healthy control (HC) based on our criteria (see **Methods**). We additionally removed five participants with MFD over 0.2mm. Nine repeated scans were excluded. Three more participants were removed due to issues with scanning sequences. A final sample of 97 HCs were included in our group comparison analysis.

### Stroop-assessed cognitive control

As described above, fMRI data from 70 participants with OUD passed quality control. Three of these individuals did not have Stroop data and were excluded from further analysis related to cognitive control. To compute the accuracy interference scores, the portion of trials where participants responded correctly was computed for both the incongruent and control conditions. Next, we subtracted the accurate portion from the incongruent and control conditions. Five participants were excluded due to having outlier performance scores (determined using MATLAB; > 3 median absolute deviations from the median). To compute the response time interference scores, we subtracted the average response time from all accurate trials in the congruent trial from the average response time from all accurate trials in the control condition. Two participants were removed since response time information during the incongruent condition was unavailable. We additionally excluded four participants with outlier response time interference scores. Accuracy and response time interference scores did not differ significantly by sex (accuracy: t(59)=0.984, p=0.329; t(59)=-0.452, p=0.653).

### Brain states identification

We replicated our prior work to identify recurring brain states. Nonlinear manifold learning and 2-step Diffusion Mapping projected task-based fMRI data from the Human Connectome Project S500 release into a low-dimensional space (2,3). We used minimally preprocessed HCP data from six different tasks (motor, working memory, social, emotional, relational, and gambling) from 390 participants (see 3 for information on quality control and exclusion criteria).

After task data were projected to the low-dimensional space, time points showing similar activity patterns were located closer together. K-means clustering then identified four recurring brain states with distinct activation patterns. The number of brain states was determined using the Calinski-Harabasz criterion (4). We characterized these brain states as fixation, high-cognition, low-cognition, and transition based on the prominent task conditions associated with these brain states (**Supplementary Table 1**). For instance, the fixation state mainly included time points from the fixation condition. The high-cognition state included time points from complex cognitive paradigms such as working memory, emotion, relational, gambling, and social. The low-cognition state involved time points from the motor task, the 0-back working memory condition, and the neutral emotion condition. Lastly, the transition state consisted of time points from the cue condition across various task paradigms. The centroid of each state cluster was extracted to serve as a representative time point in later analyses.

In our prior work (5), we investigated how canonical brain networks contributed to each of these brain states. To this end, we identified the activated and deactivated brain regions (i.e., activation above or below 0, respectively; arbitrary unit) for each representative time point in a set of canonical brain networks. The activation or deactivation percentage was next computed by dividing the number of activated or deactivated brain regions by the total number of brain regions in a network.

Different canonical networks were activated to varying extent in each brain state (**Supplementary Table 2**). However, brain network activation patterns largely followed what cognitive processes each brain state supported. For instance, the entire motor network was activated during low-cognition state whereas the high-cognition state was linked to frontoparietal network activation and default mode network deactivation.

### General linear modeling (GLM)

As an exploratory analysis, we also examined whether brain responses during the cue paradigm demonstrated an association with cognitive control. For each participant, a GLM was used to model brain activity. The design matrix included the cue regressor, mean, linear, quadratic, and cubic trend terms, and a 24-parameter motion model. The cue regressor was convolved with a standard hemodynamic response function. Resultant beta maps were spatially smoothed with a 6-mm Gaussian kernel and warped into common space. Beta coefficients were extracted from each node of the Shen-268 atlas and correlated with ACC and RT interference scores.

---

**Supplementary Figure 1.**
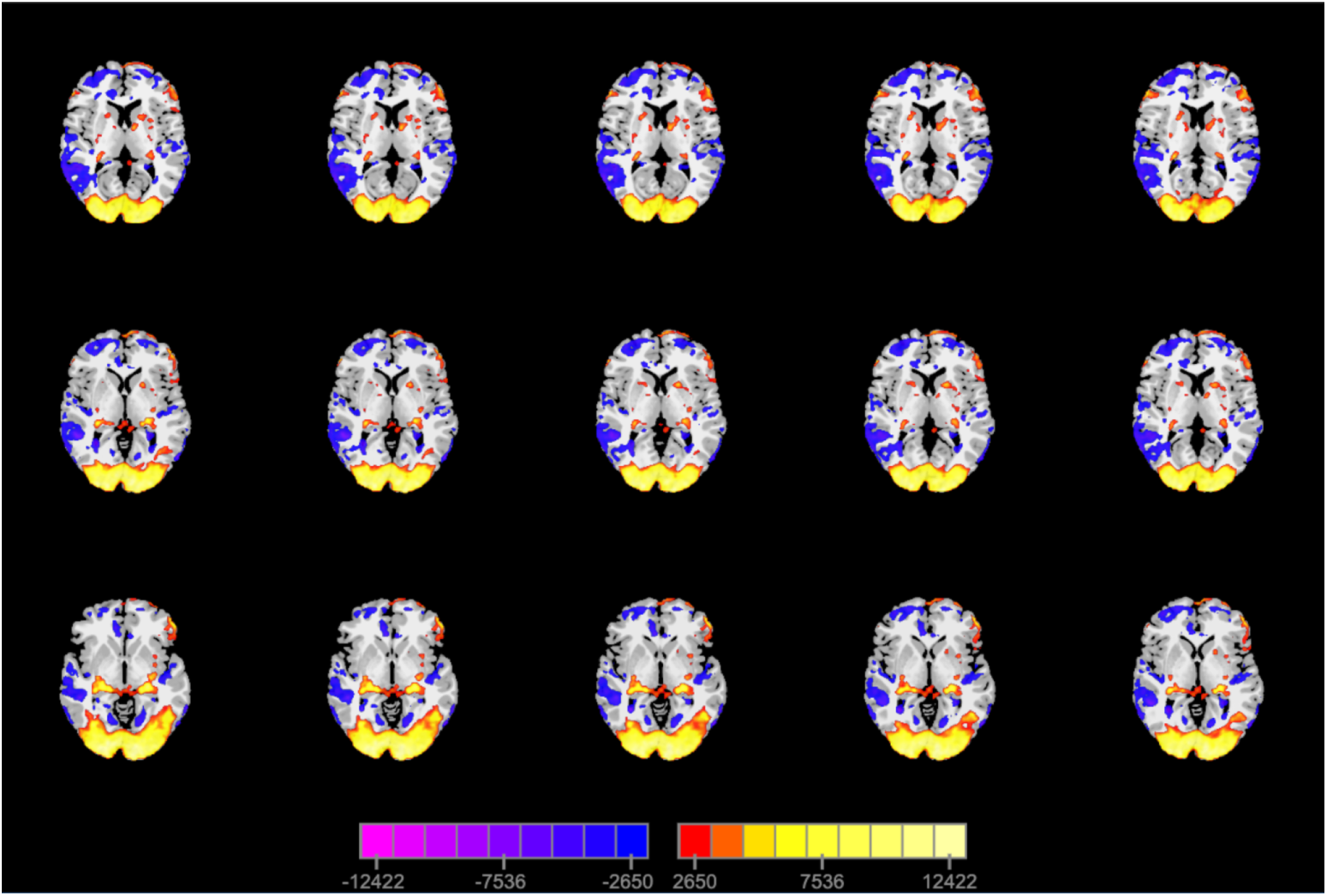
Activations during the drug cue task.

**Supplementary Table 1.** Number of volumes associated with each task condition for the four recurring brain states.

|  | Fixation | High-cognition | Low-cognition | Transition |
| --- | --- | --- | --- | --- |
| Fixation | 635 | 0 | 20 | 65 |
| Cue | 41 | 3 | 6 | 158 |
| Working memory (0 back) | 10 | 56 | 99 | 123 |
| Working memory (2 back) | 1 | 201 | 10 | 76 |
| Emotion (Fear) | 10 | 42 | 0 | 48 |
| Emotion (Neutral) | 23 | 12 | 99 | 16 |
| Gambling (Win) | 0 | 100 | 25 | 35 |
| Gambling (Loss) | 0 | 101 | 10 | 49 |
| Motor (Tongue) | 0 | 1 | 52 | 12 |
| Motor (Left foot) | 8 | 5 | 41 | 13 |
| Motor (Left hand) | 7 | 0 | 45 | 15 |
| Motor (Right foot) | 0 | 0 | 55 | 12 |
| Motor (Right hand) | 0 | 1 | 50 | 15 |
| Social (Mental) | 0 | 113 | 0 | 47 |
| Social (Random) | 0 | 111 | 0 | 109 |
| Relational (Match) | 3 | 9 | 17 | 40 |
| Relational (Relation) | 3 | 90 | 5 | 9 |

**Supplementary Table 2.** Networks showing the highest activation and deactivation percentages for each state.

|  | Fixation | High-cognition | Low-cognition | Cue/transition |
| --- | --- | --- | --- | --- |
| Activation percentages | DMN (88.89%) | VAs (100%) | Motor network (100%) | Visual I (100%) |
|  | Motor network (85.71%) | Visual II (88.87%) | MF (86.21%) | VAs (72.22%) |
|  | MF (82.76%) | FP (82.35%) | Cerebellum (84%) | Visual II (66.67%) |
| Deactivation percentages | VAs (94.44%) | Motor network (87.76%) | Visual I (100%) | MF (89.66%) |
|  | Visual I (66.67%) | DMN (83.33%) | Visual II, VAs, and DMN (66.67%) | DMN (88.87%) |
|  | Visual II (66.67%) | Subcortical (79.31%) |  | Motor (83.67%) |

This table shows the canonical functional networks with the three highest activation and deactivation percentages for each brain state. The actual activation and deactivation percentage values were included in parentheses. DMN, default mode network; MF, medial frontal network; VAs, visual association network; FP, frontoparietal network.

**Supplementary Table 3.** Individuals with opioid use disorder who reported polysubstance use.

|  | Naturalistic dataset (N=76) | Drug cue dataset (N=70) |
| --- | --- | --- |
| Alcohol | 59 | 57 |
| Amphetamines | 13 | 14 |
| Barbiturates | 5 | 4 |
| Cannabis | 69 | 61 |
| Cocaine | 67 | 63 |
| Hallucinogens | 31 | 29 |
| Inhalants | 6 | 6 |
| Other sedatives/hypnotics/tranquilizers | 20 | 21 |

**Supplementary Table 4.** State engagement variability during naturalistic stimuli ANOVA covariates.

|  | Sex | Age | Sex-by-group interaction | Age-by-group interaction |
| --- | --- | --- | --- | --- |
| Fixation | $F(1,167)=0.065$;<br>$p=0.799$ | $F(1,167)=4.202$;<br>$p=0.042$ | $F(1,167)=0.001$;<br>$p=0.974$ | $F(1,167)=5.326$;<br>$p=0.022$ |
| High-cognition | $F(1,167)=0.566$;<br>$p=0.453$ | $F(1,167)=7.173$;<br>$p=0.008$ | $F(1,167)=0.003$;<br>$p=0.959$ | $F(1,167)=4.193$;<br>$p=0.042$ |
| Low-cognition | $F(1,167)=1.070$;<br>$p=0.303$ | $F(1,167)=6.609$;<br>$p=0.011$ | $F(1,167)=0.365$;<br>$p=0.546$ | $F(1,167)=7.094$;<br>$p=0.008$ |
| Transition | $F(1,167)=0.083$;<br>$p=0.774$ | $F(1,167)=9.937$;<br>$p=0.002$ | $F(1,167)=0.234$;<br>$p=0.629$ | $F(1,167)=3.817$;<br>$p=0.052$ |

**Supplementary Table 5.** State engagement variability between rest and cue conditions.

|  | Add 1 lag to time indices | Add 2 lags to time indices |
| --- | --- | --- |
| Fixation | $t(69)=2.591$; $p=0.012$ | $t(69)=3.178$; $p=0.002$ |
| High-cognition | $t(69)=2.702$; $p=0.009$ | $t(69)=3.223$; $p=0.002$ |
| Low-cognition | $t(69)=5.526$; $p<0.001$ | $t(69)=6.482$; $p<0.001$ |
| Transition | $t(69)=2.724$; $p=0.008$ | $t(69)=3.423$; $p=0.001$ |

**Supplementary Table 6.** State engagement variability during rest condition and cognitive control.

|  | Add 1 lag to time indices |  | Add 2 lags to time indices |  |
| --- | --- | --- | --- | --- |
|  | ACC Interference | RT Interference | ACC Interference | RT Interference |
| Fixation | $\rho=-0.302$;<br>$p=0.017$ | $\rho=-0.272$;<br>$p=0.034$ | $\rho=-0.281$;<br>$p=0.027$ | $\rho=-0.233$;<br>$p=0.071$ |
| High-cognition | $\rho=-0.244$;<br>$p=0.057$ | $\rho=-0.177$;<br>$p=0.173$ | $\rho=-0.248$;<br>$p=0.052$ | $\rho=-0.157$;<br>$p=0.227$ |
| Low-cognition | $\rho=-0.089$;<br>$p=0.490$ | $\rho=-0.362$;<br>$p=0.004$ | $\rho=-0.068$;<br>$p=0.599$ | $\rho=-0.334$;<br>$p=0.009$ |
| Transition | $\rho=-0.394$;<br>$p=0.002$ | $\rho=-0.235$;<br>$p=0.069$ | $\rho=-0.383$;<br>$p=0.002$ | $\rho=-0.228$;<br>$p=0.077$ |

**Supplementary Table 7.**
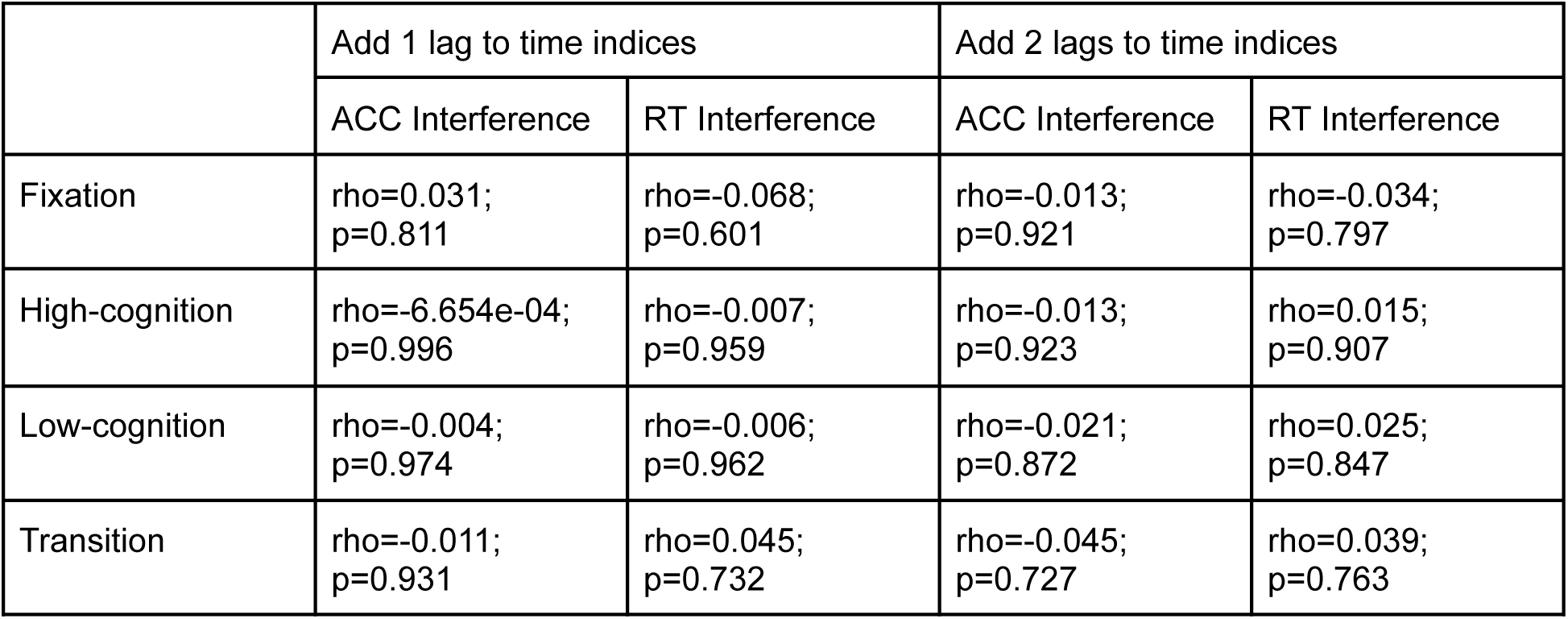
State engagement variability during cue condition and cognitive control.

**Supplementary Table 8.** Task activation during the cue paradigm and cognitive control.

| Node number | ACC Interference |  |  | RT Interference |  |  |
| --- | --- | --- | --- | --- | --- | --- |
|  | r | p | q | r | p | q |
| 1 | 0.13 | 0.33 | 0.89 | 0.19 | 0.14 | 0.99 |
| 2 | 0.05 | 0.70 | 0.89 | 0.18 | 0.16 | 0.99 |
| 3 | 0.04 | 0.75 | 0.89 | 0.13 | 0.34 | 0.99 |
| 4 | -0.07 | 0.60 | 0.89 | 0.13 | 0.33 | 0.99 |
| 5 | -0.09 | 0.49 | 0.89 | 0.05 | 0.73 | 0.99 |
| 6 | -0.08 | 0.55 | 0.89 | 0.05 | 0.67 | 0.99 |
| 7 | 0.00 | 0.97 | 0.91 | 0.13 | 0.31 | 0.99 |
| 8 | 0.05 | 0.70 | 0.89 | 0.04 | 0.73 | 0.99 |
| 9 | -0.15 | 0.23 | 0.89 | -0.02 | 0.85 | 0.99 |
| 10 | -0.09 | 0.47 | 0.89 | -0.04 | 0.76 | 0.99 |
| 11 | -0.20 | 0.11 | 0.89 | 0.10 | 0.46 | 0.99 |
| 12 | -0.05 | 0.71 | 0.89 | 0.02 | 0.86 | 0.99 |
| 13 | -0.21 | 0.10 | 0.89 | -0.09 | 0.50 | 0.99 |
| 14 | 0.02 | 0.87 | 0.90 | 0.08 | 0.53 | 0.99 |
| 15 | -0.18 | 0.16 | 0.89 | 0.00 | 0.98 | 1.00 |
| 16 | 0.05 | 0.69 | 0.89 | -0.17 | 0.20 | 0.99 |
| 17 | 0.06 | 0.62 | 0.89 | 0.22 | 0.09 | 0.99 |
| 18 | -0.01 | 0.91 | 0.90 | 0.11 | 0.38 | 0.99 |
| 19 | 0.04 | 0.74 | 0.89 | 0.06 | 0.67 | 0.99 |
| 20 | -0.09 | 0.50 | 0.89 | -0.05 | 0.70 | 0.99 |
| 21 | 0.05 | 0.72 | 0.89 | -0.02 | 0.85 | 0.99 |
| 22 | 0.21 | 0.10 | 0.89 | 0.01 | 0.95 | 1.00 |
| 23 | 0.04 | 0.75 | 0.89 | -0.07 | 0.59 | 0.99 |
| 24 | 0.09 | 0.50 | 0.89 | 0.04 | 0.75 | 0.99 |
| 25 | -0.01 | 0.93 | 0.90 | -0.06 | 0.66 | 0.99 |
| 26 | -0.05 | 0.70 | 0.89 | 0.17 | 0.18 | 0.99 |
| 27 | 0.09 | 0.48 | 0.89 | -0.01 | 0.96 | 1.00 |
| 28 | -0.15 | 0.25 | 0.89 | 0.08 | 0.55 | 0.99 |
| 29 | 0.00 | 0.99 | 0.91 | 0.22 | 0.10 | 0.99 |
| 30 | -0.14 | 0.28 | 0.89 | 0.14 | 0.30 | 0.99 |
| 31 | 0.17 | 0.18 | 0.89 | -0.04 | 0.74 | 0.99 |
| 32 | -0.11 | 0.40 | 0.89 | 0.04 | 0.76 | 0.99 |
| 33 | 0.04 | 0.73 | 0.89 | -0.06 | 0.64 | 0.99 |
| 34 | -0.07 | 0.58 | 0.89 | 0.14 | 0.29 | 0.99 |
| 35 | -0.14 | 0.27 | 0.89 | 0.06 | 0.66 | 0.99 |
| 36 | -0.13 | 0.33 | 0.89 | -0.04 | 0.76 | 0.99 |
| 37 | 0.00 | 0.98 | 0.91 | -0.05 | 0.71 | 0.99 |
| 38 | -0.19 | 0.13 | 0.89 | 0.03 | 0.81 | 0.99 |
| 39 | 0.04 | 0.78 | 0.89 | -0.07 | 0.59 | 0.99 |
| 40 | -0.05 | 0.72 | 0.89 | -0.11 | 0.41 | 0.99 |
| 41 | -0.06 | 0.63 | 0.89 | 0.09 | 0.48 | 0.99 |
| 42 | -0.19 | 0.13 | 0.89 | 0.04 | 0.77 | 0.99 |
| 43 | -0.05 | 0.73 | 0.89 | 0.12 | 0.34 | 0.99 |
| 44 | -0.05 | 0.69 | 0.89 | 0.18 | 0.16 | 0.99 |
| 45 | -0.17 | 0.18 | 0.89 | 0.00 | 0.98 | 1.00 |
| 46 | -0.13 | 0.32 | 0.89 | -0.05 | 0.72 | 0.99 |
| 47 | -0.16 | 0.21 | 0.89 | 0.20 | 0.12 | 0.99 |
| 48 | 0.09 | 0.51 | 0.89 | 0.07 | 0.58 | 0.99 |
| 49 | 0.04 | 0.79 | 0.89 | 0.13 | 0.30 | 0.99 |
| 50 | -0.02 | 0.90 | 0.90 | -0.06 | 0.62 | 0.99 |
| 51 | 0.02 | 0.88 | 0.90 | 0.00 | 0.99 | 1.00 |
| 52 | 0.00 | 0.97 | 0.91 | 0.06 | 0.63 | 0.99 |
| 53 | 0.03 | 0.82 | 0.89 | -0.06 | 0.67 | 0.99 |
| 54 | 0.00 | 0.99 | 0.91 | -0.21 | 0.11 | 0.99 |
| 55 | 0.09 | 0.48 | 0.89 | 0.12 | 0.36 | 0.99 |
| 56 | -0.05 | 0.72 | 0.89 | 0.10 | 0.44 | 0.99 |
| 57 | -0.08 | 0.51 | 0.89 | 0.04 | 0.74 | 0.99 |
| 58 | -0.12 | 0.35 | 0.89 | -0.02 | 0.85 | 0.99 |
| 59 | -0.05 | 0.70 | 0.89 | -0.03 | 0.84 | 0.99 |
| 60 | -0.12 | 0.35 | 0.89 | -0.04 | 0.76 | 0.99 |
| 61 | 0.03 | 0.80 | 0.89 | -0.04 | 0.78 | 0.99 |
| 62 | -0.05 | 0.68 | 0.89 | -0.15 | 0.24 | 0.99 |
| 63 | -0.04 | 0.76 | 0.89 | -0.10 | 0.46 | 0.99 |
| 64 | -0.09 | 0.51 | 0.89 | -0.10 | 0.45 | 0.99 |
| 65 | 0.05 | 0.71 | 0.89 | -0.14 | 0.30 | 0.99 |
| 66 | 0.03 | 0.79 | 0.89 | -0.02 | 0.86 | 0.99 |
| 67 | 0.22 | 0.09 | 0.89 | -0.04 | 0.75 | 0.99 |
| 68 | -0.16 | 0.20 | 0.89 | -0.13 | 0.32 | 0.99 |
| 69 | 0.15 | 0.24 | 0.89 | 0.00 | 0.99 | 1.00 |
| 70 | 0.06 | 0.62 | 0.89 | 0.03 | 0.82 | 0.99 |
| 71 | 0.02 | 0.90 | 0.90 | -0.03 | 0.84 | 0.99 |
| 72 | 0.05 | 0.70 | 0.89 | 0.02 | 0.86 | 0.99 |
| 73 | 0.06 | 0.63 | 0.89 | -0.01 | 0.92 | 1.00 |
| 74 | -0.01 | 0.92 | 0.90 | -0.12 | 0.35 | 0.99 |
| 75 | 0.00 | 0.99 | 0.91 | 0.12 | 0.37 | 0.99 |
| 76 | 0.13 | 0.32 | 0.89 | -0.08 | 0.55 | 0.99 |
| 77 | -0.12 | 0.34 | 0.89 | -0.04 | 0.77 | 0.99 |
| 78 | 0.28 | 0.03 | 0.89 | 0.03 | 0.83 | 0.99 |
| 79 | 0.14 | 0.27 | 0.89 | 0.02 | 0.85 | 0.99 |
| 80 | 0.02 | 0.86 | 0.90 | 0.14 | 0.29 | 0.99 |
| 81 | 0.18 | 0.17 | 0.89 | 0.12 | 0.37 | 0.99 |
| 82 | 0.05 | 0.68 | 0.89 | 0.00 | 0.99 | 1.00 |
| 83 | -0.17 | 0.19 | 0.89 | -0.01 | 0.93 | 1.00 |
| 84 | -0.14 | 0.27 | 0.89 | 0.00 | 0.99 | 1.00 |
| 85 | -0.04 | 0.79 | 0.89 | 0.06 | 0.65 | 0.99 |
| 86 | -0.13 | 0.30 | 0.89 | -0.08 | 0.55 | 0.99 |
| 87 | -0.05 | 0.71 | 0.89 | -0.08 | 0.54 | 0.99 |
| 88 | -0.13 | 0.30 | 0.89 | 0.07 | 0.58 | 0.99 |
| 89 | -0.12 | 0.34 | 0.89 | -0.12 | 0.36 | 0.99 |
| 90 | -0.15 | 0.24 | 0.89 | 0.06 | 0.67 | 0.99 |
| 91 | -0.15 | 0.25 | 0.89 | 0.14 | 0.28 | 0.99 |
| 92 | -0.06 | 0.67 | 0.89 | 0.18 | 0.16 | 0.99 |
| 93 | -0.30 | 0.02 | 0.89 | 0.00 | 1.00 | 1.00 |
| 94 | 0.03 | 0.81 | 0.89 | 0.17 | 0.19 | 0.99 |
| 95 | -0.24 | 0.06 | 0.89 | -0.01 | 0.95 | 1.00 |
| 96 | -0.12 | 0.34 | 0.89 | -0.10 | 0.43 | 0.99 |
| 97 | -0.04 | 0.78 | 0.89 | 0.14 | 0.27 | 0.99 |
| 98 | -0.08 | 0.56 | 0.89 | 0.00 | 0.98 | 1.00 |
| 99 | -0.01 | 0.96 | 0.91 | 0.14 | 0.28 | 0.99 |
| 100 | 0.07 | 0.61 | 0.89 | 0.03 | 0.83 | 0.99 |
| 101 | -0.26 | 0.04 | 0.89 | 0.09 | 0.51 | 0.99 |
| 102 | 0.05 | 0.72 | 0.89 | -0.03 | 0.85 | 0.99 |
| 103 | -0.19 | 0.15 | 0.89 | 0.02 | 0.91 | 1.00 |
| 104 | -0.03 | 0.79 | 0.89 | 0.07 | 0.60 | 0.99 |
| 105 | -0.12 | 0.37 | 0.89 | 0.10 | 0.46 | 0.99 |
| 106 | -0.08 | 0.54 | 0.89 | 0.08 | 0.52 | 0.99 |
| 107 | -0.04 | 0.76 | 0.89 | 0.31 | 0.02 | 0.99 |
| 108 | -0.10 | 0.44 | 0.89 | 0.10 | 0.46 | 0.99 |
| 109 | -0.11 | 0.37 | 0.89 | 0.02 | 0.91 | 1.00 |
| 110 | -0.17 | 0.19 | 0.89 | 0.02 | 0.90 | 1.00 |
| 111 | 0.05 | 0.69 | 0.89 | 0.09 | 0.50 | 0.99 |
| 112 | -0.03 | 0.82 | 0.89 | 0.09 | 0.51 | 0.99 |
| 113 | -0.13 | 0.31 | 0.89 | 0.01 | 0.92 | 1.00 |
| 114 | 0.05 | 0.70 | 0.89 | 0.11 | 0.41 | 0.99 |
| 115 | -0.11 | 0.40 | 0.89 | 0.10 | 0.46 | 0.99 |
| 116 | -0.12 | 0.34 | 0.89 | 0.06 | 0.63 | 0.99 |
| 117 | -0.20 | 0.13 | 0.89 | -0.01 | 0.93 | 1.00 |
| 118 | -0.10 | 0.43 | 0.89 | 0.21 | 0.11 | 0.99 |
| 119 | -0.08 | 0.52 | 0.89 | 0.07 | 0.61 | 0.99 |
| 120 | -0.01 | 0.95 | 0.91 | 0.03 | 0.83 | 0.99 |
| 121 | -0.04 | 0.73 | 0.89 | 0.00 | 0.98 | 1.00 |
| 122 | -0.11 | 0.41 | 0.89 | 0.00 | 1.00 | 1.00 |
| 123 | 0.04 | 0.77 | 0.89 | 0.16 | 0.22 | 0.99 |
| 124 | -0.05 | 0.71 | 0.89 | -0.14 | 0.27 | 0.99 |
| 125 | -0.04 | 0.76 | 0.89 | 0.08 | 0.55 | 0.99 |
| 126 | -0.13 | 0.30 | 0.89 | 0.13 | 0.32 | 0.99 |
| 127 | -0.04 | 0.77 | 0.89 | 0.14 | 0.27 | 0.99 |
| 128 | -0.06 | 0.63 | 0.89 | 0.07 | 0.57 | 0.99 |
| 129 | 0.10 | 0.44 | 0.89 | 0.19 | 0.15 | 0.99 |
| 130 | 0.08 | 0.53 | 0.89 | 0.08 | 0.56 | 0.99 |
| 131 | -0.12 | 0.34 | 0.89 | -0.08 | 0.53 | 0.99 |
| 132 | -0.24 | 0.06 | 0.89 | 0.02 | 0.88 | 1.00 |
| 133 | -0.17 | 0.19 | 0.89 | -0.05 | 0.69 | 0.99 |
| 134 | -0.01 | 0.91 | 0.90 | 0.14 | 0.29 | 0.99 |
| 135 | -0.03 | 0.83 | 0.90 | 0.04 | 0.77 | 0.99 |
| 136 | 0.23 | 0.07 | 0.89 | -0.07 | 0.59 | 0.99 |
| 137 | 0.05 | 0.69 | 0.89 | -0.08 | 0.56 | 0.99 |
| 138 | -0.05 | 0.68 | 0.89 | 0.02 | 0.88 | 0.99 |
| 139 | 0.05 | 0.69 | 0.89 | -0.04 | 0.77 | 0.99 |
| 140 | -0.07 | 0.57 | 0.89 | -0.01 | 0.92 | 1.00 |
| 141 | -0.12 | 0.34 | 0.89 | -0.03 | 0.83 | 0.99 |
| 142 | -0.09 | 0.49 | 0.89 | 0.01 | 0.94 | 1.00 |
| 143 | 0.06 | 0.62 | 0.89 | -0.04 | 0.76 | 0.99 |
| 144 | -0.13 | 0.33 | 0.89 | 0.06 | 0.63 | 0.99 |
| 145 | -0.18 | 0.16 | 0.89 | -0.10 | 0.43 | 0.99 |
| 146 | -0.02 | 0.89 | 0.90 | -0.05 | 0.70 | 0.99 |
| 147 | 0.11 | 0.38 | 0.89 | -0.09 | 0.50 | 0.99 |
| 148 | -0.10 | 0.46 | 0.89 | -0.19 | 0.15 | 0.99 |
| 149 | -0.06 | 0.62 | 0.89 | -0.11 | 0.41 | 0.99 |
| 150 | -0.11 | 0.38 | 0.89 | 0.04 | 0.78 | 0.99 |
| 151 | 0.14 | 0.27 | 0.89 | -0.08 | 0.54 | 0.99 |
| 152 | 0.04 | 0.77 | 0.89 | 0.11 | 0.41 | 0.99 |
| 153 | -0.04 | 0.76 | 0.89 | 0.06 | 0.62 | 0.99 |
| 154 | 0.14 | 0.27 | 0.89 | 0.15 | 0.23 | 0.99 |
| 155 | -0.09 | 0.48 | 0.89 | -0.01 | 0.94 | 1.00 |
| 156 | 0.03 | 0.84 | 0.90 | -0.01 | 0.96 | 1.00 |
| 157 | 0.14 | 0.29 | 0.89 | -0.04 | 0.78 | 0.99 |
| 158 | 0.10 | 0.45 | 0.89 | -0.05 | 0.68 | 0.99 |
| 159 | 0.07 | 0.61 | 0.89 | -0.04 | 0.79 | 0.99 |
| 160 | -0.08 | 0.54 | 0.89 | 0.09 | 0.51 | 0.99 |
| 161 | -0.17 | 0.18 | 0.89 | -0.05 | 0.68 | 0.99 |
| 162 | -0.11 | 0.38 | 0.89 | 0.05 | 0.71 | 0.99 |
| 163 | 0.03 | 0.82 | 0.89 | 0.08 | 0.54 | 0.99 |
| 164 | -0.13 | 0.30 | 0.89 | 0.11 | 0.39 | 0.99 |
| 165 | 0.06 | 0.67 | 0.89 | -0.02 | 0.86 | 0.99 |
| 166 | -0.05 | 0.67 | 0.89 | -0.04 | 0.74 | 0.99 |
| 167 | -0.02 | 0.88 | 0.90 | 0.00 | 1.00 | 1.00 |
| 168 | -0.03 | 0.80 | 0.89 | 0.13 | 0.34 | 0.99 |
| 169 | -0.01 | 0.92 | 0.90 | 0.07 | 0.57 | 0.99 |
| 170 | -0.05 | 0.71 | 0.89 | -0.21 | 0.10 | 0.99 |
| 171 | 0.02 | 0.87 | 0.90 | -0.04 | 0.78 | 0.99 |
| 172 | -0.06 | 0.62 | 0.89 | -0.04 | 0.77 | 0.99 |
| 173 | -0.05 | 0.72 | 0.89 | -0.20 | 0.13 | 0.99 |
| 174 | 0.00 | 1.00 | 0.91 | 0.05 | 0.70 | 0.99 |
| 175 | -0.01 | 0.93 | 0.90 | 0.07 | 0.61 | 0.99 |
| 176 | -0.20 | 0.12 | 0.89 | -0.03 | 0.83 | 0.99 |
| 177 | 0.11 | 0.39 | 0.89 | -0.07 | 0.60 | 0.99 |
| 178 | 0.00 | 1.00 | 0.91 | 0.20 | 0.12 | 0.99 |
| 179 | -0.03 | 0.85 | 0.90 | 0.02 | 0.87 | 0.99 |
| 180 | -0.11 | 0.40 | 0.89 | -0.09 | 0.49 | 0.99 |
| 181 | -0.21 | 0.10 | 0.89 | -0.04 | 0.77 | 0.99 |
| 182 | -0.06 | 0.66 | 0.89 | -0.03 | 0.82 | 0.99 |
| 183 | -0.02 | 0.91 | 0.90 | -0.16 | 0.22 | 0.99 |
| 184 | -0.08 | 0.55 | 0.89 | 0.17 | 0.20 | 0.99 |
| 185 | -0.02 | 0.90 | 0.90 | -0.18 | 0.16 | 0.99 |
| 186 | -0.04 | 0.74 | 0.89 | -0.04 | 0.76 | 0.99 |
| 187 | -0.07 | 0.59 | 0.89 | 0.02 | 0.85 | 0.99 |
| 188 | 0.00 | 0.99 | 0.91 | 0.01 | 0.96 | 1.00 |
| 189 | 0.08 | 0.56 | 0.89 | 0.11 | 0.38 | 0.99 |
| 190 | -0.02 | 0.90 | 0.90 | -0.05 | 0.69 | 0.99 |
| 191 | 0.00 | 0.99 | 0.91 | -0.08 | 0.52 | 0.99 |
| 192 | 0.04 | 0.73 | 0.89 | -0.17 | 0.19 | 0.99 |
| 193 | 0.02 | 0.87 | 0.90 | 0.03 | 0.83 | 0.99 |
| 194 | -0.17 | 0.18 | 0.89 | -0.01 | 0.91 | 1.00 |
| 195 | -0.15 | 0.26 | 0.89 | 0.03 | 0.79 | 0.99 |
| 196 | -0.20 | 0.11 | 0.89 | 0.06 | 0.62 | 0.99 |
| 197 | -0.08 | 0.53 | 0.89 | -0.04 | 0.75 | 0.99 |
| 198 | 0.09 | 0.48 | 0.89 | -0.04 | 0.74 | 0.99 |
| 199 | 0.04 | 0.75 | 0.89 | 0.07 | 0.61 | 0.99 |
| 200 | 0.09 | 0.49 | 0.89 | -0.04 | 0.73 | 0.99 |
| 201 | -0.03 | 0.80 | 0.89 | 0.01 | 0.95 | 1.00 |
| 202 | 0.01 | 0.92 | 0.90 | -0.06 | 0.63 | 0.99 |
| 203 | 0.03 | 0.80 | 0.89 | -0.15 | 0.25 | 0.99 |
| 204 | 0.03 | 0.81 | 0.89 | -0.05 | 0.72 | 0.99 |
| 205 | -0.09 | 0.49 | 0.89 | -0.09 | 0.49 | 0.99 |
| 206 | 0.14 | 0.28 | 0.89 | -0.02 | 0.89 | 1.00 |
| 207 | 0.09 | 0.46 | 0.89 | 0.03 | 0.85 | 0.99 |
| 208 | 0.02 | 0.90 | 0.90 | 0.11 | 0.38 | 0.99 |
| 209 | 0.04 | 0.75 | 0.89 | -0.16 | 0.21 | 0.99 |
| 210 | 0.10 | 0.45 | 0.89 | -0.15 | 0.24 | 0.99 |
| 211 | 0.13 | 0.33 | 0.89 | -0.02 | 0.85 | 0.99 |
| 212 | 0.27 | 0.03 | 0.89 | 0.09 | 0.48 | 0.99 |
| 213 | 0.02 | 0.89 | 0.90 | 0.03 | 0.80 | 0.99 |
| 214 | 0.11 | 0.42 | 0.89 | 0.05 | 0.68 | 0.99 |
| 215 | 0.07 | 0.57 | 0.89 | -0.05 | 0.72 | 0.99 |
| 216 | -0.07 | 0.60 | 0.89 | -0.04 | 0.79 | 0.99 |
| 217 | -0.21 | 0.10 | 0.89 | 0.08 | 0.53 | 0.99 |
| 218 | -0.18 | 0.15 | 0.89 | -0.10 | 0.43 | 0.99 |
| 219 | -0.19 | 0.14 | 0.89 | -0.04 | 0.76 | 0.99 |
| 220 | -0.15 | 0.24 | 0.89 | -0.11 | 0.38 | 0.99 |
| 221 | -0.17 | 0.19 | 0.89 | 0.04 | 0.78 | 0.99 |
| 222 | -0.07 | 0.61 | 0.89 | -0.22 | 0.09 | 0.99 |
| 223 | -0.07 | 0.58 | 0.89 | 0.03 | 0.82 | 0.99 |
| 224 | -0.21 | 0.09 | 0.89 | -0.04 | 0.77 | 0.99 |
| 225 | -0.13 | 0.31 | 0.89 | 0.03 | 0.82 | 0.99 |
| 226 | -0.19 | 0.15 | 0.89 | 0.12 | 0.36 | 0.99 |
| 227 | -0.09 | 0.48 | 0.89 | -0.05 | 0.72 | 0.99 |
| 228 | 0.00 | 0.98 | 0.91 | 0.17 | 0.20 | 0.99 |
| 229 | 0.07 | 0.62 | 0.89 | 0.03 | 0.80 | 0.99 |
| 230 | -0.01 | 0.91 | 0.90 | 0.08 | 0.55 | 0.99 |
| 231 | -0.04 | 0.77 | 0.89 | 0.21 | 0.10 | 0.99 |
| 232 | 0.22 | 0.09 | 0.89 | 0.04 | 0.75 | 0.99 |
| 233 | 0.07 | 0.58 | 0.89 | 0.09 | 0.52 | 0.99 |
| 234 | -0.02 | 0.89 | 0.90 | -0.16 | 0.23 | 0.99 |
| 235 | -0.10 | 0.46 | 0.89 | 0.10 | 0.46 | 0.99 |
| 236 | -0.07 | 0.58 | 0.89 | 0.13 | 0.32 | 0.99 |
| 237 | -0.15 | 0.23 | 0.89 | 0.03 | 0.80 | 0.99 |
| 238 | -0.06 | 0.66 | 0.89 | 0.10 | 0.45 | 0.99 |
| 239 | -0.22 | 0.09 | 0.89 | 0.17 | 0.20 | 0.99 |
| 240 | -0.07 | 0.56 | 0.89 | 0.10 | 0.43 | 0.99 |
| 241 | 0.08 | 0.56 | 0.89 | 0.24 | 0.07 | 0.99 |
| 242 | -0.01 | 0.94 | 0.91 | 0.09 | 0.47 | 0.99 |
| 243 | -0.01 | 0.95 | 0.91 | 0.05 | 0.70 | 0.99 |
| 244 | -0.16 | 0.22 | 0.89 | 0.16 | 0.22 | 0.99 |
| 245 | -0.18 | 0.16 | 0.89 | 0.02 | 0.86 | 0.99 |
| 246 | 0.01 | 0.92 | 0.90 | 0.15 | 0.24 | 0.99 |
| 247 | -0.07 | 0.59 | 0.89 | 0.08 | 0.53 | 0.99 |
| 248 | -0.04 | 0.76 | 0.89 | 0.09 | 0.51 | 0.99 |
| 249 | -0.03 | 0.82 | 0.89 | 0.15 | 0.26 | 0.99 |
| 250 | -0.17 | 0.18 | 0.89 | -0.02 | 0.85 | 0.99 |
| 251 | -0.06 | 0.67 | 0.89 | 0.18 | 0.16 | 0.99 |
| 252 | 0.05 | 0.70 | 0.89 | 0.28 | 0.03 | 0.99 |
| 253 | 0.04 | 0.75 | 0.89 | 0.09 | 0.47 | 0.99 |
| 254 | -0.09 | 0.47 | 0.89 | 0.08 | 0.56 | 0.99 |
| 255 | -0.25 | 0.05 | 0.89 | 0.00 | 0.99 | 1.00 |
| 256 | 0.04 | 0.77 | 0.89 | 0.07 | 0.57 | 0.99 |
| 257 | -0.03 | 0.81 | 0.89 | 0.24 | 0.06 | 0.99 |
| 258 | 0.10 | 0.42 | 0.89 | 0.24 | 0.06 | 0.99 |
| 259 | -0.08 | 0.53 | 0.89 | 0.11 | 0.38 | 0.99 |
| 260 | -0.03 | 0.84 | 0.90 | 0.04 | 0.79 | 0.99 |
| 261 | 0.03 | 0.79 | 0.89 | -0.12 | 0.34 | 0.99 |
| 262 | 0.11 | 0.39 | 0.89 | 0.05 | 0.68 | 0.99 |
| 263 | -0.11 | 0.41 | 0.89 | 0.05 | 0.69 | 0.99 |
| 264 | -0.05 | 0.72 | 0.89 | 0.12 | 0.37 | 0.99 |
| 265 | -0.29 | 0.02 | 0.89 | 0.01 | 0.92 | 1.00 |
| 266 | 0.04 | 0.75 | 0.89 | 0.13 | 0.34 | 0.99 |
| 267 | -0.18 | 0.16 | 0.89 | -0.07 | 0.60 | 0.99 |
| 268 | 0.08 | 0.54 | 0.89 | 0.09 | 0.51 | 0.99 |

**Supplementary Table 9.** Associations between cognitive control and state engagement variability during the first fixation block (i.e., before opioid-related stimulus presentations)

|  | Accuracy Interference | Response Time Interference |
| --- | --- | --- |
| Fixation | $\rho=-0.024$; $p=0.852$ | $\rho=-0.206$; $p=0.112$ |
| High-cognition | $\rho=-0.026$; $p=0.841$ | $\rho=-0.094$; $p=0.470$ |
| Low-cognition | $\rho=-0.068$; $p=0.601$ | $\rho=-0.099$; $p=0.446$ |
| Transition | $\rho=-0.042$; $p=0.749$ | $\rho=0.221$; $p=0.087$ |

